# REMINDer randomized controlled study: Feasibility and impact of an online multimodal mind-body intervention in older adults

**DOI:** 10.1101/2024.11.10.24317080

**Authors:** Miranka Wirth, Selina Stamer, Odile Podolski, Annemarie Maßalsky, Sabine C. Koch, Olga Klimecki

## Abstract

**Background:** The increase in life expectancy and age-related diseases, such as Alzheimer’s disease, is a global challenge. Given the drawbacks of pharmacological treatments, it is important to pursue non-pharmacological strategies for dementia risk reduction. To effectively promote health and well-being in later life, multimodal, low-threshold, and cost-effective lifestyle interventions are needed.

**Methods:** *REMINDer* is a monocentric, outcome assessor-blinded, randomized controlled pilot study to assess the feasibility and impact of an online (home-based and live-streamed) multimodal intervention. The 6-week (two one-hour sessions/week) mind-body group intervention will be compared to a 6-week passive control (waitlist with delayed intervention) using a cross-over (AB-BA) design. The intervention was designed for older adults and includes music, dance-based movement, and mindfulness. A total of *N*=50 cognitively unimpaired older adults will be enrolled and randomized into the two intervention arms using a block randomization with a 1:1 allocation ratio.

**Results:** Outcomes will be assessed at pre-intervention, post-intervention, and follow-up using digital assessments of online questionnaires. Primary outcomes include feasibility, operationalized by adherence rates, and preliminary effectiveness of the intervention. The latter will be assessed by changes in self-reported mental and physical well-being, operationalized by the Short-Form Health Survey (SF-12). Secondary outcomes will include changes in cognitive, motor, sensory, emotional/affective, social, and lifestyle health behaviors.

**Discussion:** The study will provide evidence of the feasibility and preliminary effectiveness of an online multimodal mind-body intervention based on “Arts for Health” activities. If successful, the study may inform accessible lifestyle strategies to improve mental health and well-being and other risk factors for dementia in older adults.

**Trail Registration:** ClinicalTrials.gov, NCT06530277

**Protocol:** SR-EK-477112023

## 1 Introduction

### 1.1 Research background

Age-related neurodegenerative diseases, including Alzheimer’s disease (AD), are increasing rapidly and represent a significant public health and societal challenge. Cases of dementia, with AD being the most prevalent type, are expected to triple by 2050, placing a high burden on patients, caregivers, and society (1). Even though there is progress regarding disease-modifying therapies, it is important to pursue non-pharmacological options for the prevention and early intervention of dementia. A large proportion of the risk factors for developing AD are related to potentially modifiable lifestyle factors, including physical, cognitive, psychological and social factors (2). Low-threshold, accessible, and cost-efficient lifestyle interventions are needed to effectively reduce the adverse effects of aging and prevent AD in the long term.

#### 1.1.1 Multimodal interventions: Rational and motivation

To achieve long-term health benefits in older adults and eventually prevent AD, it could be most effective to implement intervention strategies that target multiple risk factors at once (3). Such multimodal or multidomain lifestyle interventions are considered a promising strategy to counteract the impact of age- and disease-related adversities on brain and cognitive health (4, 5). In animal models, environmental enrichment that combines motor, cognitive, sensory, and social stimulation has been shown to have beneficial effects including hippocampal neurogenesis, synaptogenesis, and expression of brain-derived neurotrophic factor compared to low-enrichment (standard housing) (6-8). However, translating animal models incorporating an enriched environment to human behavior and lifestyle is challenging (7).

Improving the mental health and well-being of older adults must be important goals of lifestyle interventions. Psychological features or symptoms, including anxiety, depression, and loneliness, have been identified as risk factors for the development/progression of cognitive impairment and dementia (9-11) and often manifest in the pre-symptomatic stages of AD (12). In this line, greater emphasize needs to be given to prevention strategies that incorporate lifestyle activities providing “*depth, purpose, meaning, and social embedding*” to achieve healthy aging of body and mind (13, 14). It is anticipated that holistic or integrated lifestyle activities involving an "*embodied mind in motion*" (15, 16) may enhance reserve and resilience of older adults to address the health challenges associated with aging and age-related disease.

#### 1.1.2 Dance: A multi-domain health facilitator

A possible "naturalistic" translation of the "*embodied mind in motion*" is offered by “Arts for Health” activities related music and dance (13, 16). Dancing has been associated with a reduced risk for developing AD (17) and provides an inherent and simultaneous multimodal – motor, cognitive, sensory, emotional, and social – integration, engaging large-scale brain networks involved in action, perception, emotion and cognition (18-20). As such, dance movement interventions have been proposed as effective tools for the embodied prevention and intervention of diverse populations, including older adults at risk of or living with dementia (19, 21-23). Moreover, multimodal enrichment strategies are expected to have a higher ecological validity (24) and elicit far transfer effects (24, 25) in humans.

Indeed, there is emerging evidence that dance movement interventions, including various dance types, dance aerobics, and/or dance/movement therapy (DMT), can promote the health and well-being across multiple domains in older adults with normal and/or impaired cognition. These include improvements in cognitive (26-29), motor/physical (30, 31), and psychological/mental (22, 26, 32, 33) functioning – even for shorter-term interventions of 12-weeks or less (26, 34). However, the existing evidence on the mental health benefits in older adults without dementia is still sparse and inconclusive (22, 33, 35). Given the contribution of psychological risk/health factors to healthy mental ageing and dementia risk reduction, further research is needed to determine the effectiveness of dance movement interventions in this context.

### 1.2 *REMINDer*: An online multimodal mind-body intervention

As a result of the existing research, we have recently developed and documented a multimodal mind-body group intervention, called ***REMIND (“An Envi<u>r</u>onmental <u>E</u>nrich<u>m</u>ent Interve<u>n</u>tion to Prevent <u>D</u>ementia”)***. This intervention was specifically designed for older adults and is available as an open-access manual (36). The *REMIND* program uses an integrated or combined approach that aims to promote the health and well-being of older adults across multiple cognitive, psychological, physical, and social domains. In brief, the intervention integrates three core components “*music, movement, and mind*” in order to combine music with dance-based movement and mindfulness. Each of these core components has beneficial effects on a wide range of health outcomes, including improved brain and mental/cognitive health, in older adults without dementia. This is evidenced by a large body of studies from our and others groups for music (e.g., 37, 38, 39), dance movement (e.g., 22, 40, 41), and mindfulness (e.g., 42, 43, 44). In the *REMIND* program, the three core components are practiced in combination resp. simultaneously to generate a multimodal – motor, cognitive, sensory, emotional, and social – enrichment and facilitate an "*embodied mind in motion*" (13, 16).

In the present study, we will conduct the *REMIND* program as an online (home-based and live-streamed) intervention, thereafter termed ***REMINDer* (*REMIND - <u>e</u>lectronic t<u>r</u>ial*)**, adapted to the needs/challenges of the older target population. To effectively reach older people from diverse ethnic, economic, and geographical backgrounds, including those in rural and underserved areas, access to and delivery of interventions must be as low-threshold and accessible as possible (45). Previously, multimodal mind-body group interventions, including dance, mindfulness-based DMT, and Tai Chi, have been delivered online (via videoconferencing) to healthy older participants (46-48) and older participants with cognitive (49, 50) and physical (51, 52) conditions. Online interventions (also known as teleinterventions) have been shown to be feasible, well-accepted, safe, and preliminary effective in older adults (45, 53) – comparable to in-person interventions (54).

## 2 Objective of this study

Given the importance of psychological risk factors in aging processes and the development of AD (2), more research using high-quality randomized controlled studies is needed to assess the impact of multimodal mind-body interventions on the mental health and well-being of older adults. Specifically, there is an urgent need to evaluate changes across multiple health domains, including psychological, physical, and social well-being, and quality of life in response to mind-body interventions. This may facilitate their low-threshold and accessible implementation as embodied prevention and/or early intervention tools in diverse older populations (21-23).

We will perform a randomized controlled pilot study with an online multimodal mind-body group intervention in cognitively unimpaired older adults. The 6-week (two one-hour sessions/week) intervention program (*REMINDer*) will be compared to a 6-week passive control condition (waitlist with delayed intervention) using a cross-over (AB-BA) design. The intervention is delivered as an interactive group program in the online setting (home-based and live-streamed) via a licensed videoconferencing software. The primary objective of the *REMINDer* study is to evaluate the feasibility and preliminary effectiveness of the online multimodal intervention. The feasibility of the intervention will be assessed by adherence rates. The impact of the intervention will be assessed by mental and physical well-being as operationalized by the Short-Form Health Survey (SF-12) (55, 56). In addition, we will assess the effects of the intervention on secondary health outcomes, including self-reported cognitive, motor, sensory, emotional, social, and lifestyle behaviors.

## 3 Methods

### 3.1 Study design and setting

The *REMINDer* study is a monocentric, randomized controlled pilot study with blinded outcome assessment conducted at the German Center for Neurodegenerative Diseases (DZNE) Dresden, Dresden, Germany. The recruitment area is Saxony and Thuringia in particular, and Germany in general. Based on previous research (34, 57) and feasibility considerations, this pilot study will include a shorter-term intervention duration of 6 weeks with a moderate frequency of 2 sessions per week and 60 min duration per session, conducted as an AB-BA crossover study design with the following intervention sequences:

1. Sequence: Beginning with the online multimodal intervention (*REMINDer)* and then transitioning to a no-intervention phase (sequence: AB).
2. Sequence: Starting with a no-intervention phase and subsequently moving to the online multimodal intervention (*REMINDer)* (sequence: BA).

### 3.2 Participants

A random sample of cognitively unimpaired older adults will be recruited from the general community in Germany through written and oral announcements (i.e., online and newspaper articles) and flyers about the study. The list of inclusion and exclusion criteria is provided in **Table 1**. Participants will provide written informed consent during study enrollment. Participants will be informed that they can discontinue participation at any time during the study period without negative consequences.

**Table 1.** Eligibility criteria.

| Inclusion criteria | Exclusion criteria |
| --- | --- |
| Retired older adults (aged > 60 years*)<br>Fluency in German language<br>Cognitively unimpaired, as assessed via the Six-Item Screener (SIS) $\geq 4$ points (out of 6)<br>Access to a wireless internet connection and a digital device with speakers, screen, and camera at home<br>Access to a personal e-mail address<br>Sufficient in-home space<br>Availability throughout the study and its measurement time points<br>Ability to provide written informed consent | Regular rigorous physical exercise as defined by regular aerobic exercise (> 1.5 hours/week) during the last 6 months<br>Regular prior experience (> 1 hour/week) with Tai Chi, dance movements, or other mind-body practices during the last 6 months<br>Disabilities that limit participation in online moment programs: Presence of physical ineligibility or mobility restrictions through a history of falls, not being able to walk, reliance on a walking aid etc.<br>Severe uncorrected auditory or visual impairments that limit the ability to listen to online instructions or to observe online movements<br>Diagnosis of cognitive impairment and any type of dementia<br>Diagnosis of motor disorders (e.g., Parkinson's disease, multiple sclerosis)<br>History of cerebral disease (e.g., brain tumor)<br>Severe neurological disorders (e.g., epilepsy)<br>Diagnosis of psychiatric disorders (e.g., depression)<br>Other chronic medical disorders that limit physical activity (e.g., advanced cardiac or respiratory disease, severe hypertension)<br>Substance abuse (excessive smoking, alcohol consumption, drug abuse)<br>Current participation in another research study<br>(Note: Subclinical comorbidities of psychological symptoms will not lead to exclusion.) |
| * Note: An earlier version of the study protocol included older adults aged 60 – 80 years. |  |

\*\*\*\*\*

### 3.3 Assessment of study measures

#### 3.3.1 Participant timeline

The *REMINDer* study will include the following phases for each participant: study enrollment (t0), a baseline/pre-intervention assessment (t1), followed by a 6-week intervention or no-intervention period, and a post-period assessment (t2).

Subsequently, participants will transition to the next 6-week period (either moving from intervention to no-intervention or vice versa), concluding with a follow-up assessment after 6 weeks (t3).

For the AB group ("intervention, no-intervention") t1 serves as the baseline for the intervention and t2 concludes the intervention and t3 is the follow-up. For the BA group ("no-intervention, intervention") t1 serves as the baseline for the no-intervention period, t2 concludes the no-intervention period and t3 concludes the intervention. Study flow and participant timeline are provided in **Figure 1**.

**Figure 1.**
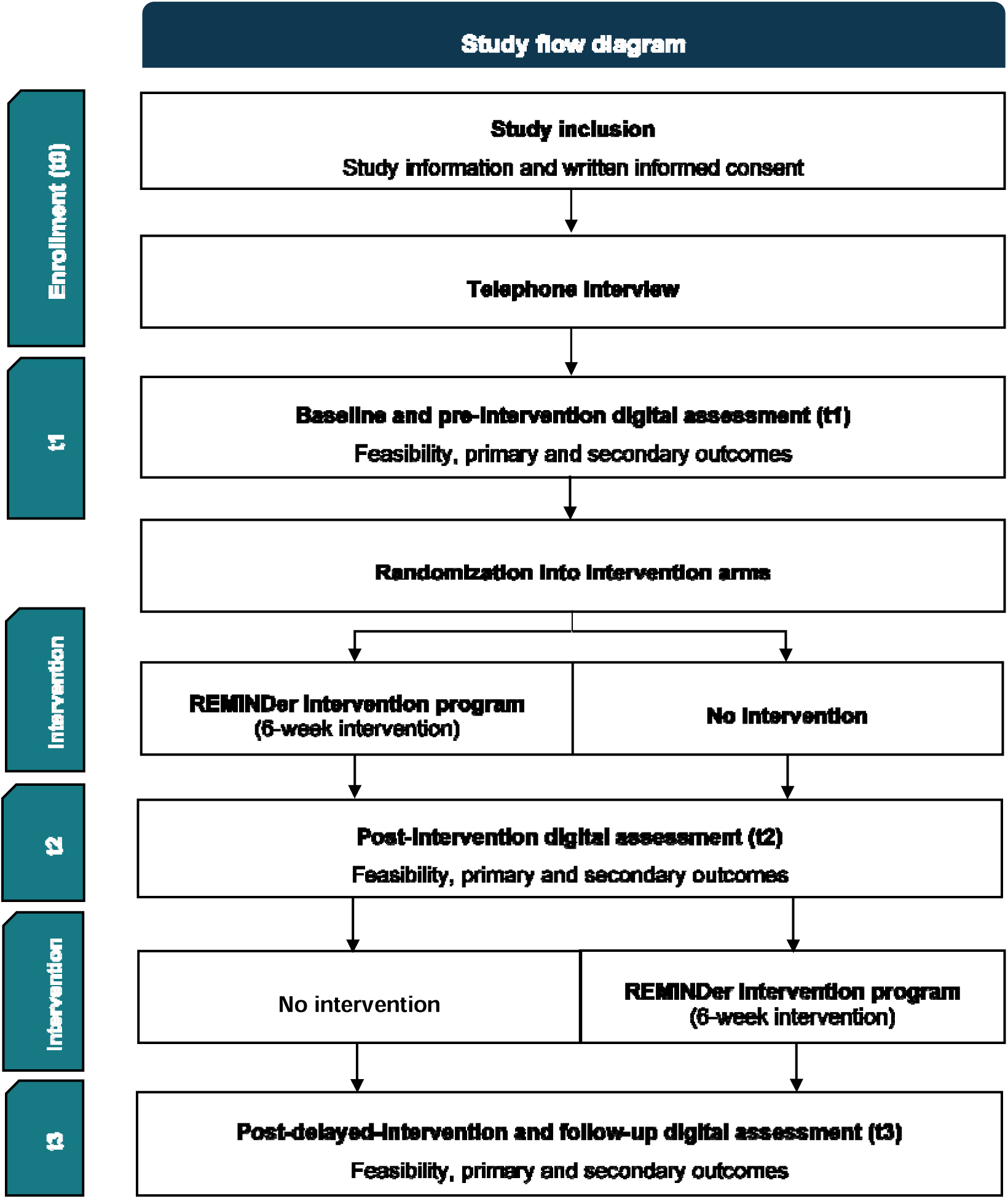
Flowchart of the *REMINDer* study.

#### 3.3.2 Data collection

A comprehensive battery of validated behavioral questionnaires will be assessed online at all stages (for details, see **Table 2).** Note that existing German versions of the questionnaires are implemented. Additional measures will be obtained at baseline to characterize participants and identify potential confounders. The outcome measures will be obtained using digital assessments via the LimeSurvey software in agreement with data protection regulations. Assessments take approximately 60 minutes (per assessment/time point). Assessments will be provided in sections of 20 minutes on 3 consecutive days to reduce the workload. An additional 20 minutes are expected for the baseline assessment as well as about 20 minutes for the telephone screening. Assessments of study measures are detailed in **Table 2**. Possible health risks and measures to avoid them are described in the section **Ethics, safety, and monitoring**. Note that the outcome assessment will take place online; no visit to the study institution is required at any time point.

**Table 2:**
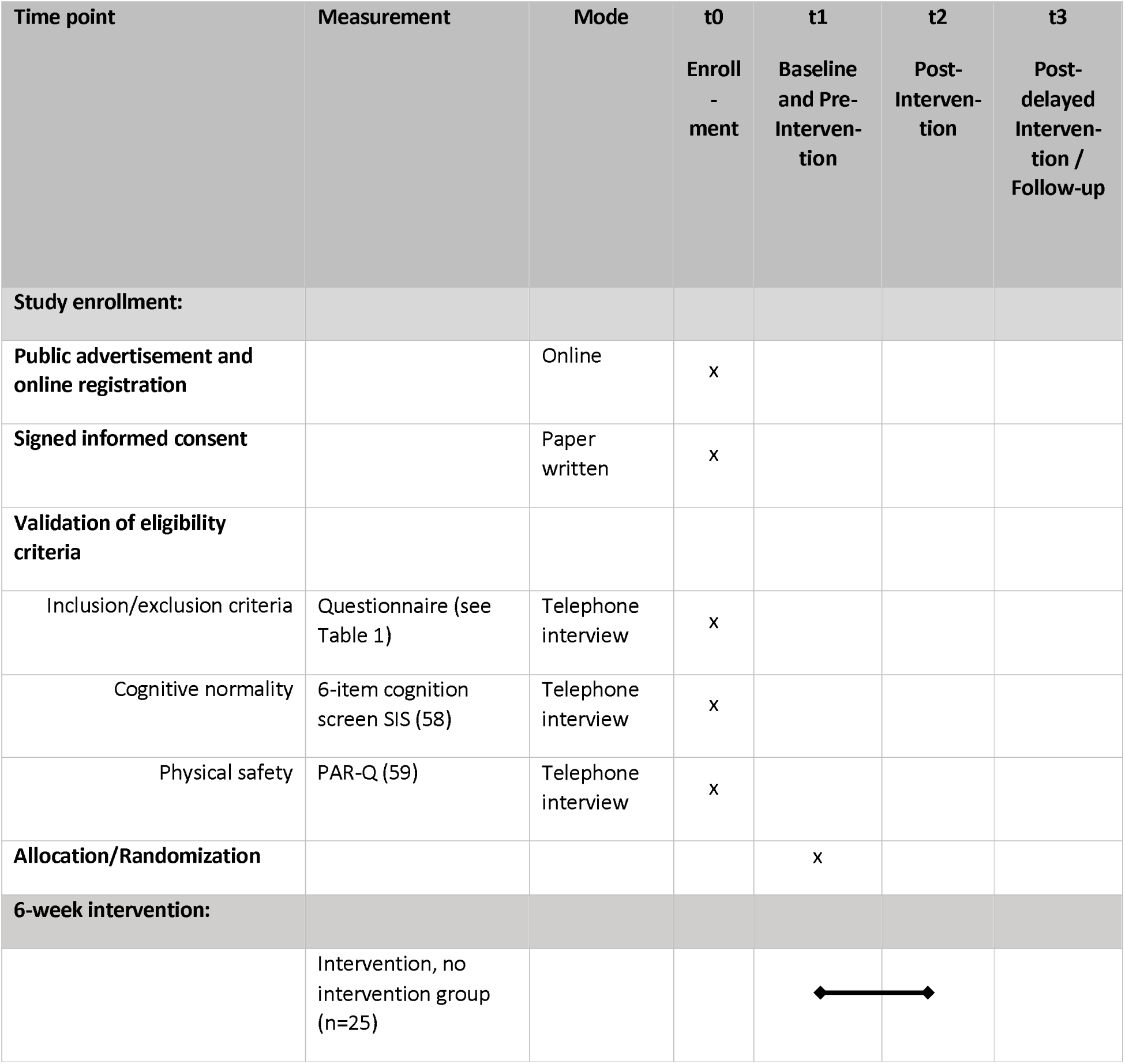
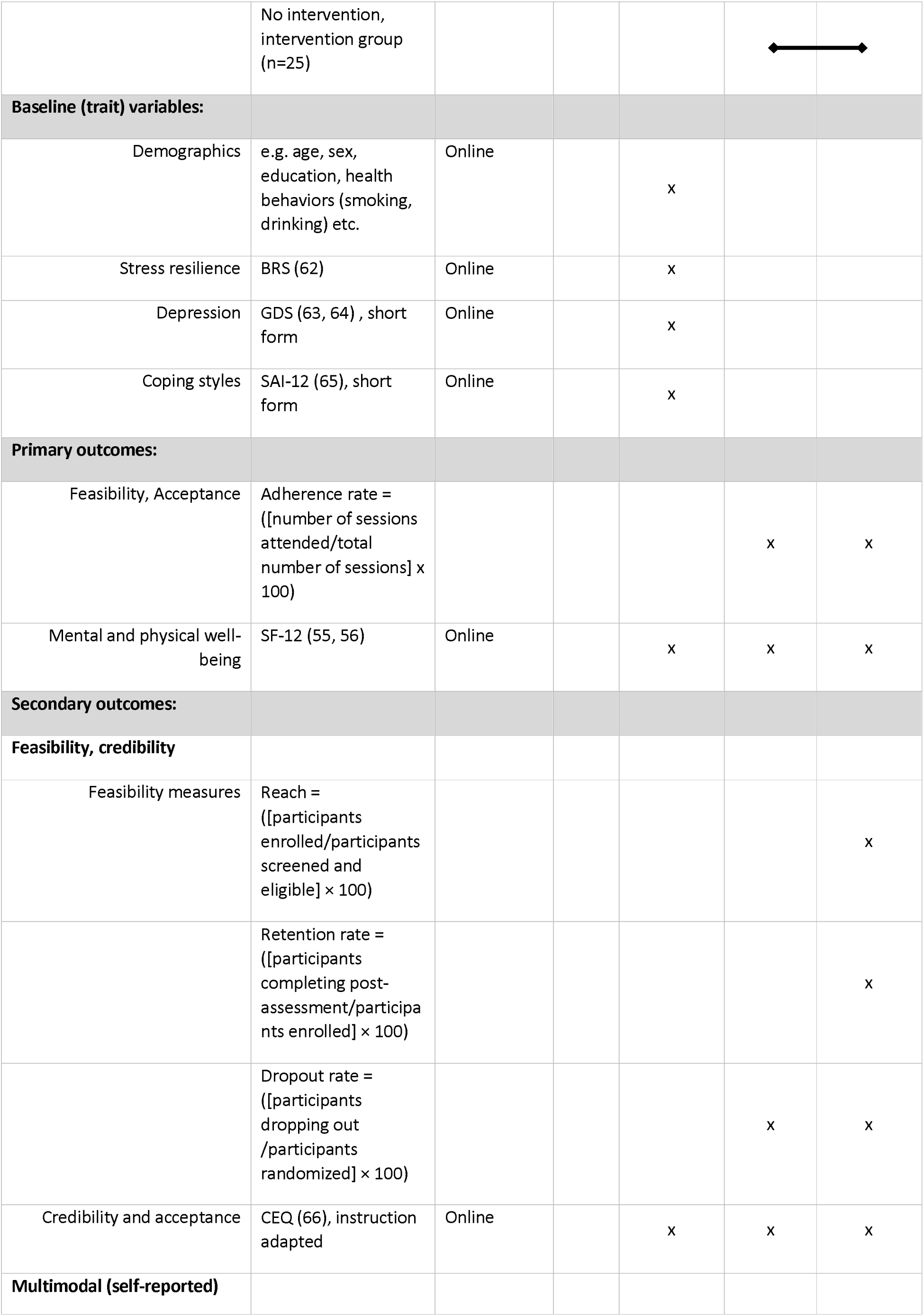
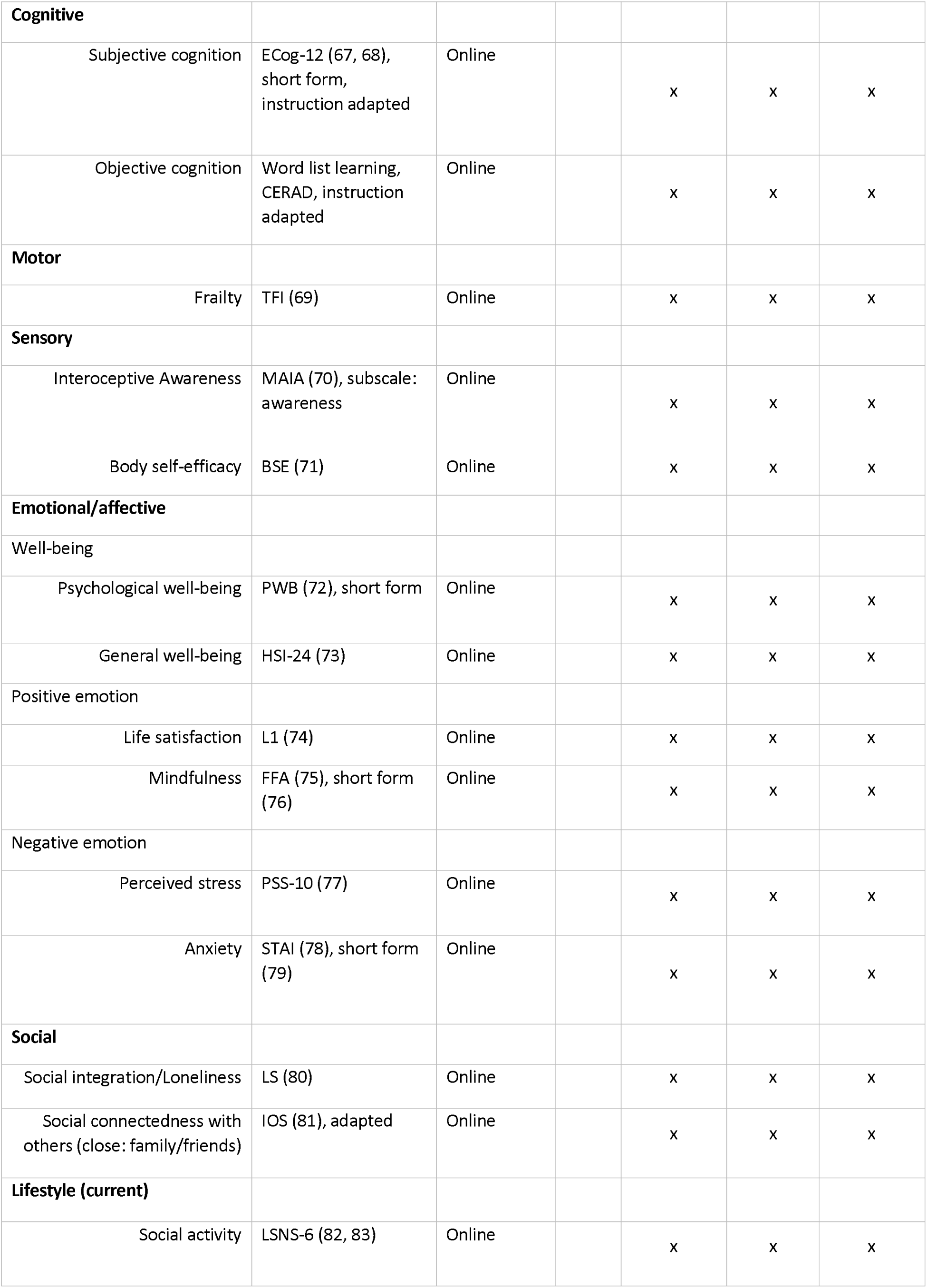
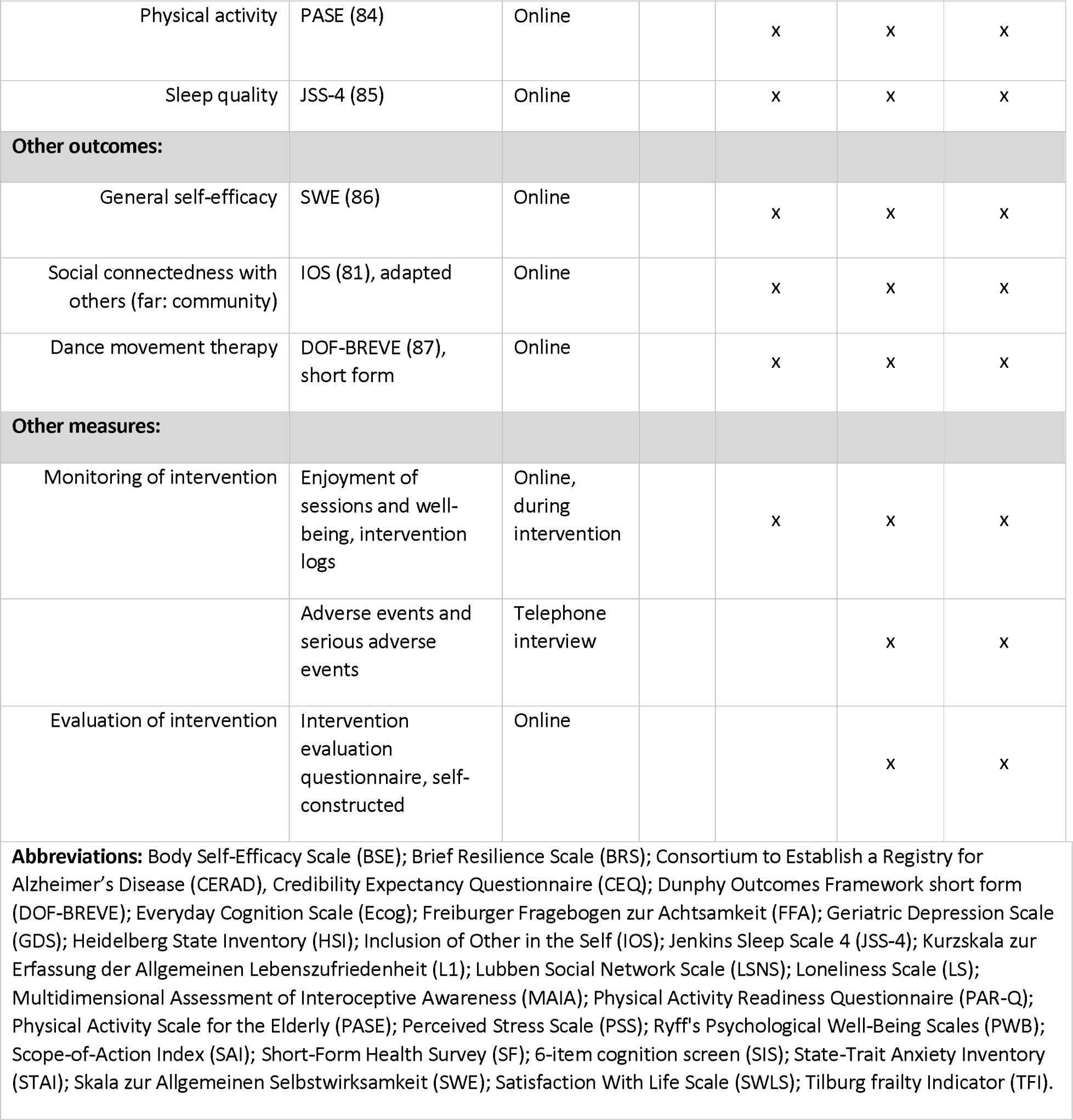
R*E*MINDer study assessments.

#### 3.3.3 Study visits

##### Enrollment (v0)

The enrollment procedure for the online *REMINDer* study will include the following steps. First, public advertisements for the study including eligibility criteria will be placed. Individuals interested in participating in the study will visit the registration website (via URL or QR-Code) and register contact information online in order to receive detailed study information. Second, each registered person will receive information about the study content and eligibility criteria, together with a written informed consent form, by email or by post. Persons interested in study participation will sign the consent form and return it to the study team. The consent form clearly states that participation in the *REMINDer* study requires the study personnel to validate the eligibility criteria during a telephone interview (see below).

##### Telephone interview (v0)

After written informed consent is obtained, participants will be assessed by telephone interview according to the eligibility criteria listed in **Table 1**. The interview will take approximately 20 minutes and includes standardized questions related to medical, cognitive, and physical eligibility validation. Medical conditions are assessed by questions related to serious physical, neurological, and/or psychiatric disorders, and substance abuse. Cognitive normality is assessed by the Six-Item Screener (SIS). A person is eligible to participate in the study if at least 4 out of 6 items are scored (58). The Physical Activity Readiness Questionnaire (PAR-Q) (59) is administered to assess potential physical constraints. If one or more questions on the PAR-Q are answered "yes", the individual will be asked to seek medical advice and provide confirmation that the current health status meets the requirements for participation in the study. If, during the telephone interview, it becomes apparent that the person meets exclusion criteria that preclude study participation, the respondent will be excluded from the study.

Note that during study enrollment, participants will have the possibility to clarify any unanswered questions about participating in the *REMINDer study* via email or phone. Additionally, Standard Operating Procedures (SOPs) on setting up and handling the videoconferencing platform will be issued. Participants will then be randomized to the intervention arms according to the procedure described in **section Randomization and Blinding** and will proceed to the baseline assessment.

##### Baseline/pre-intervention (t1)

Baseline and pre-intervention assessment will take place in close temporal proximity before the start of the intervention. In the baseline assessment, data on detailed demographic and psychometric characteristics that may influence or moderate the outcome of the intervention are collected. In the pre-intervention assessment, all primary and secondary outcomes are collected using established and valid questionnaires (**Table 2**).

##### Post-intervention (t2) and Post-delayed intervention and Follow-up (t3)

The assessments are similar to the pre-intervention (t1) assessment. As shown in **Table 2**, the digital assessments (t1, t2, and t3) are almost identical for both study arms, except for the evaluation of the intervention, which is provided after completion of the *REMINDer* program. The baseline variable survey is omitted at t2 and t3.

In addition, participants will be monitored at the end of each intervention week for their subjective well-being using an adapted version of the World Health Organization-Five Well-Being Index (WHO-5) (60, 61).

#### 3.3.4 Randomization and blinding

Participants will be randomly assigned to one of the two intervention arms using block randomization stratified by age and sex using a computer-based algorithm. We aim to enroll a total of *n* = 25 participants in the AB group, and *n* = 25 participants in the BA group, that is, *N* = 50 participants in total. Note that spouses may be allowed in the same group. After being randomly assigned to the intervention arms, participants will receive details on the intervention from the study personnel. Study outcomes will be collected and assessed using digital questionnaire batteries and by a designated statistician blinded to the group allocation. Blinding will not be broken until the last participant has been assessed at the post-delayed intervention/follow-up assessment (t3) and the analysis of the primary outcome has been completed.

### 3.4 Intervention

#### 3.4.1 Description of intervention

In the *REMINDer* study, the participants will be offered an online multimodal mind-body group intervention. The intervention is based on the *REMIND* program that was previously developed for older adults by our research group in cooperation with experts in dance and dance/movement therapy (88). A short introduction and overview of the intervention is provided below.

The overarching goal of the *REMIND* program is to activate and strengthen the brain/cognitive reserve and resilience of older adults, which can be defined as the capacity of the brain to withstand the effects of aging and the onset of dementia. (89, 90). To this aim, the intervention uses a multimodal enrichment strategy, consisting of the three core components "*music, movement, and mind*" (see **Table 3**). These core components are practiced simultaneously to generate a complex multimodal motor, cognitive, sensory, emotional and social stimulation or enrichment (see **Table 4**). Through the distinct synthesis of perception, action, and reflection, the intervention is intended to promote the health and well-being across multiple (mental, cognitive, physical and social) domains. *REMIND* constitutes a tool to promote an "*embodied mind in motion*" and embodied prevention in older adults.

**Table 3.** Core components of the *REMIND* Program.

|  | Rationale |
| --- | --- |
| **General** | The *REMIND* program combines and integrates the three core components of "*music, movement, and mind*" in order to maintain mental health and well-being in the sense of an inherent balance and synergy of the body, brain and mind. The simultaneous integration of music, movement, and mind activities and the inherent multimodal stimulation represents a key strategy in *REMIND*. The core components work synergistically together to facilitate the growth/development and preservation/maintenance of motor skills, cognitive abilities, sensory perceptions, emotions and moods within an interactive social environment. |
| **Movement** | Dance-based movement is one core component of the *REMIND* program. Most of the exercises involve dance movements practiced with mindfulness and awareness. Dance is a physical activity that provides complex stimulation to the brain by engaging a variety of cognitive processes, including spatial orientation, coordination, balance, posture and flexibility. Older adults in particular benefit from motor skill learning, both in terms of physical and mental well-being. The synergistic interaction of body and mind and the promotion of embodied movement is perceived as an essential element of mental and physical health. |
| **Music** | The second core component of the *REMIND* program is the perception and interpretation of music. The program facilitates conscious and attentive listening to and experiencing of music. The impulses of melody and rhythm are translated into movement in a creative manner under the guidance of the facilitator/trainer. The combination and integration of music listening and immediate movement is an effective approach for older individuals. Listening to music activates large-scale networks of specialized brain regions that are responsible for attention, memory, motor function, |
|  | and emotional processing. Listening to music further motivates people to move, which in turn improves their emotions and moods, as well as their physical strength, stability, and mobility. Music plays an important role in mental, physical, and social health in older adults. |
| **Mind** | The third core component of the *REMIND* program is the experience of the present internal and external environment, which we refer to and understand as mental or mind activity. The intentional practice of mindfulness plays an important role in promoting well-being in older adults. While existing exercise programs often prioritize the improvement of physical strength and performance, *REMIND* emphasizes the conscious perception, introspection, awareness, and internalization of movement, including both sensory and emotional processes. This “mind activity” can help increase mental strength and well-being, which are key elements of healthy aging. |

\*\*\*\*\*

**Table 4.**
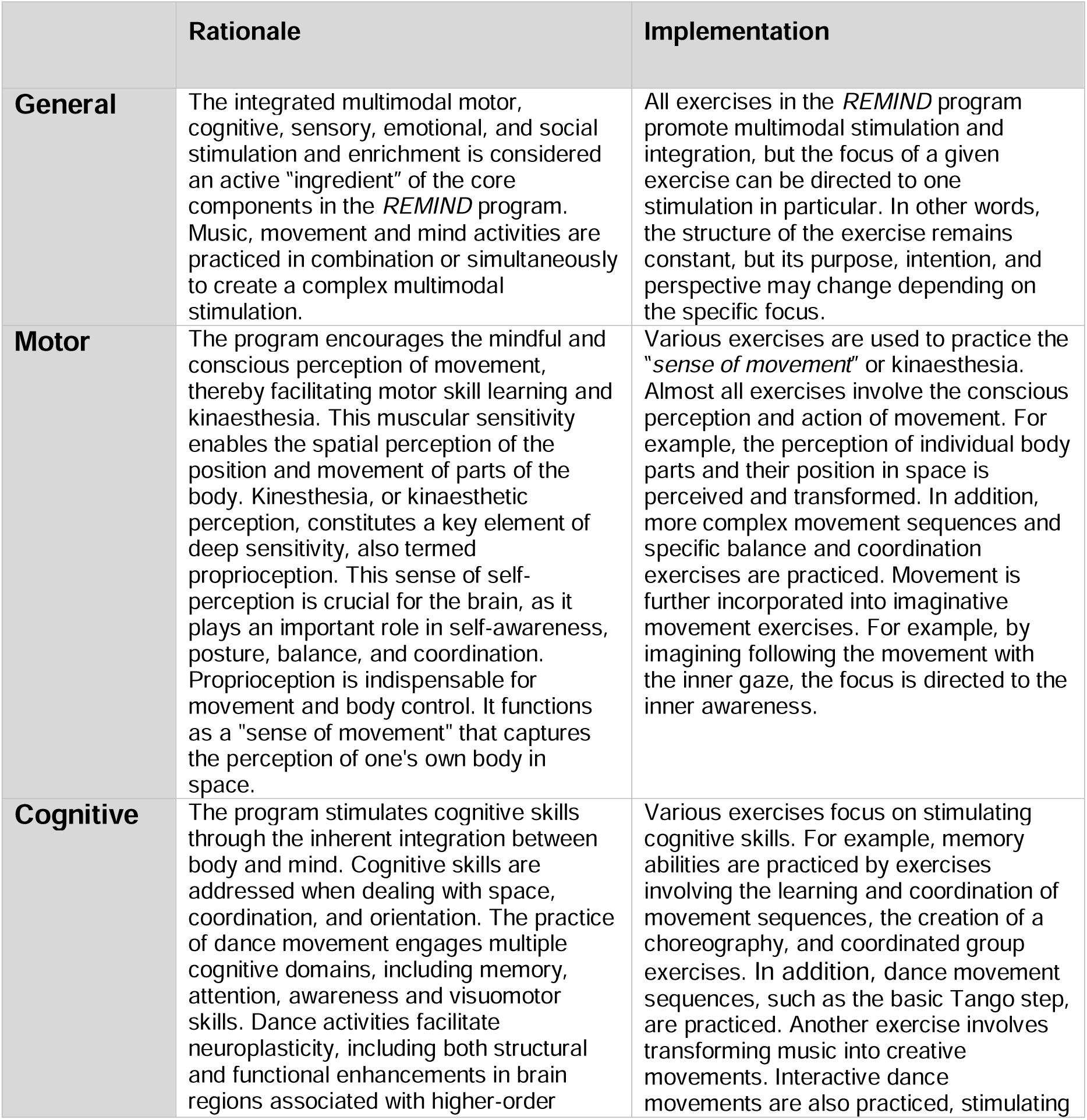
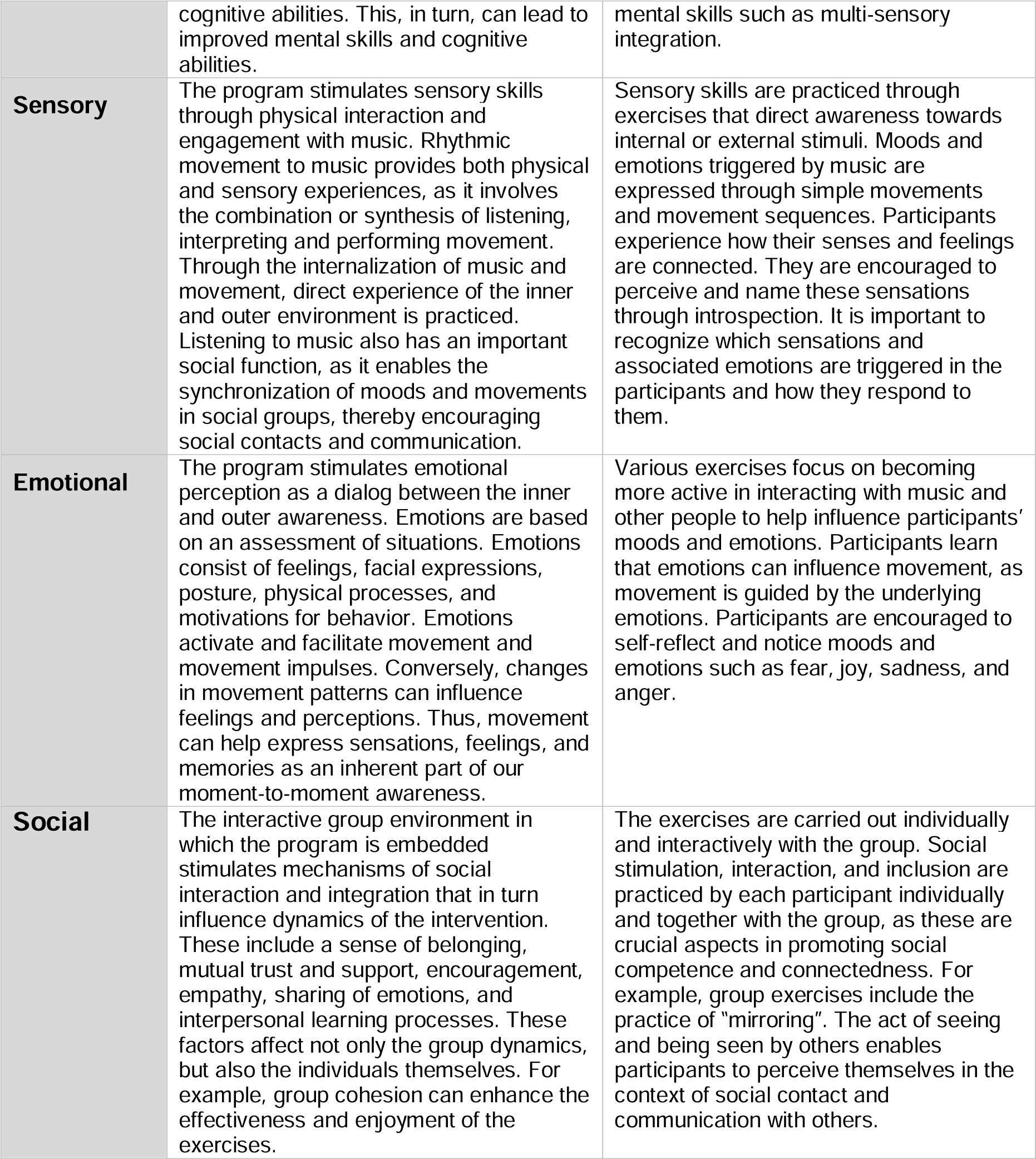
Multimodal stimulation in the *REMIND* Program.

More specifically, the *REMIND* program incorporates principles and techniques from dance/movement therapy, dance/tango teaching using the "*Sistema Dinzel*" method, and mindfulness practice. As such, the intervention offers guided movement exercises to train movement awareness, balance, and coordination. Participants are guided to listen to and experience music by translating melody and rhythm into movement. Mindfulness exercises are used to train awareness of one’s own body, perception of movement, as well as empathy. All exercises are designed to promote positive learning experiences, creativity, and self-confidence/self-efficacy. Social interaction is a key stimulation in *REMIND.* Therefore, the group completes the entire program together to enable social interaction and social integration, associated with life satisfaction, well-being, and better brain and cognitive health in older adults (e.g., 91, 92).

#### 3.4.2 Description of online delivery

For the present study, the *REMIND* program (88) is adapted to and delivered as an online (home-based and live-streamed) intervention, called *REMINDer,* The intervention will be delivered in an interactive online group environment through the implementation of a licensed videoconferencing system in compliance with data protection regulations. To ensure adequate social interaction opportunities and participants’ safety in the online environment, the intervention group will be divided into two smaller groups of about equal size. The participants will attend the live-streamed intervention sessions from their home using their own technical devices. Participants will receive access information and meeting reminders via email to increase adherence. They will be familiarized with the software environment using SOPs provided after study enrollment. Technical support will be provided to the participants throughout the intervention.

The intervention will be delivered in 2 online sessions à 60 min per week over a duration of 6 weeks. Two trained facilitators with educational backgrounds and certifications in dance/movement therapy and mindfulness practice, respectively, will guide the online intervention with exercises presented and performed in the live-streamed online environment. The program requires no previous experience. Most of the exercises are performed while sitting or standing and involve light to moderate physical activity. Single training sessions are divided into three phases: a welcome/warm-up (∼10 min), training (∼40 min), and relaxation/interactive feedback (∼10 min) phase. During each online session, participants will perform exercises that combine and integrate music, movement, and mind activities based on a well-structured manual (88).

#### 3.4.3 Choice of comparator

The comparator of this pilot study is a passive control group (waitlist with delayed intervention). Participants will be randomized to either the *REMINDer* group or the delayed-intervention group. During the waiting period, the participants engage in their normal daily lifestyle. An overview of the intervention groups is displayed in **Figure 1**.

In the AB-BA study design, the participants in the "intervention, no-intervention" (AB) group will undergo the 6-week online multimodal mind-body intervention (*REMINDer*) between t1 and t2, and then obtain no intervention between t2 and t3. Between t2 and t3 participants can decide for themselves to what extent they wish to continue the practices. Conversely, the participants in the "no-intervention, intervention" (BA) group will continue with their normal routine during the initial 6-week period between t1 and t2. They will then participate in the online multimodal intervention (*REMINDer*) during the subsequent 6-week period between t2 and t3.

### 3.5 Study outcomes

#### 3.5.1 Primary outcome

The aim of this pilot study is to determine feasibility and preliminary effectiveness of the *REMINDer* program. **Feasibility** will be assessed via adherence rates (see **Table 2**). Adherence to the intervention per participant is defined as the number of sessions attended in relation to the total number of sessions. The change from baseline/pre-intervention to post-intervention in mental and physical well-being will be assessed as the co-primary outcome. This will be evaluated using the Short-Form Health Survey (SF-12) (Ware et al., 1996; Wirtz et al., 2018), which is a self-report questionnaire with twelve items on mental and physical health outcomes.

#### 3.5.2 Secondary outcome

A detailed overview of secondary outcome measures is provided in **Figure 2**. Secondary outcomes include credibility and further measures concerning the feasibility of the intervention including reach, retention rate, and dropout rate as defined in **Table 2**. Since the control group (waitlist with delayed intervention) will receive the intervention between t2 and t3, the cross-over design will allow for the evaluation of feasibility measures at different time points. In addition, changes from pre-intervention (t1) to post-intervention (t2) will be assessed using self-reported multimodal cognitive, motor, sensory, emotional, social and lifestyle behaviors as provided in **Table 2**.

#### 3.5.3 Other outcomes

Other outcome measures include assessment of self-efficacy, closeness to community, and selected dance/movement therapy outcomes. In addition, we will monitor the intervention using enjoyment of session logs (for participants) and intervention logs (for trainers). Finally, the *REMINDer* intervention will be evaluated by the participants at the end of the intervention using an adapted evaluation questionnaire.

### 3.6 Statistical considerations

#### 3.6.1 Sample size

The pilot sample size was estimated based on the objective of assessing the feasibility of the intervention. Sample size estimation was conducted using a one-sided test for proportion against the fixed proportion of 60% with an effect size of 30% (g = 0.30). The significance level was set to 5%. The effect should be detectable with a statistical power of at least 80%. Based on these criteria, the minimum sample size was estimated to 14 participants per intervention arm (calculated in G*Power, version 3.1) (93). With an expected ‘dropout’ rate of 20%, a total of 17 participants per intervention arm was calculated. In order to increase statistical power and demonstrate an effect of the intervention, we aim to include 25 participants per intervention arm (in total: *N* = 50).This sample size exceeds the recommended sample size for pilot studies (94).

#### 3.6.2 Data analysis

The statistical analysis will be carried out using statistical software packages available in R (R Core Team, 95) Statistical analyses of the primary outcomes will be performed by a designated statistician. Statistical analyses of secondary outcomes and additional exploratory analyses will be performed by the study investigators. Data analysis will be conducted in cooperation with and under supervision of the Institute for Medical Informatics and Biometry (IMB), Technische Universität Dresden, Dresden, Germany.

##### Baseline characteristics

Baseline characteristics of participants will be summarized and compared using independent t-tests for continuous variables and a Χ^2^ test for categorical variables. Distribution of baseline characteristics between groups will be assessed to confirm a successful randomization process.

##### Analysis of primary outcomes

The primary statistical objective of this pilot study is to evaluate feasibility and preliminary effectiveness of the intervention. The primary outcome is feasibility, which will be assessed via adherence rates. A one-sided test for proportion will be used to assess whether a minimum adherence rate of 60% of participants per intervention group is achieved, although an adherence rate of at least 80% can be expected (22). The co-primary outcome is the change in mental and physical well-being as operationalized with the SF-12 (55, 56). Primary and co-primary outcomes will be assessed between t1 and t2 using an ‘intention-to-treat’ (ITT) approach, evaluating all participants randomized into the study. Note that spouses will be allowed in the same group and statistical analyses will be adapted accordingly.

##### Analysis strategy

Given the AB-BA crossover design, we will conduct a series of analyses. In a first step, we will perform between-group comparisons of primary and secondary outcomes after the completion of the first intervention period from t1 to t2. Specifically, changes in outcomes from pre-intervention (t1) to post-intervention (t2) from the AB intervention group will be contrasted with changes in outcomes from pre-intervention (t1) to post-intervention (t2) from the BA no-intervention group. Additionally, within- and between-group intervention effects will be explored including the post-delayed intervention or follow-up (t3) assessment. For example, we will assess, whether a similar pattern of changes as from t1 to t2 in the AB intervention group will be present from t2 to t3 in the BA delayed intervention group. Finally, we will explore potential moderation effects of pre-existing psychological traits on the outcomes of the intervention.

The statistical analysis strategy of this study will be informed by our previous randomized controlled studies (44, 96, 97). In particular, linear mixed models will be employed to assess changes in primary and secondary outcomes from t1 to t2 as a function of the intervention arms. For each outcome, estimates with 95% confidence intervals (CI) will be reported for the slope of change within each intervention group and the differences between the intervention groups. Effect sizes for the evaluated changes will be represented by Cohen’s d (98), estimated using the calculated mean changes. Models will be adjusted for potential confounders; in addition, unadjusted models will be explored. The primary outcome and co-primary outcome will both be evaluated at a two-sided significance level of alpha = 0.05.

To handle missing data, we will assess whether the data is missing completely at random (MCAR). Should the MCAR assumption be confirmed, the full information maximum likelihood estimation (FIML) method will be employed in order to optimize the utilization of the available data without introducing bias. Note that an exploratory analysis of the longer-term effects, from t1 to t3, will be conducted similar to the primary and co-primary outcome analysis. Overall, the statistical analysis approach will ensure a nuanced understanding of both the immediate and longer-term impacts of the intervention across both sequences (AB and BA).

## 4 Ethics, safety, and monitoring

The *REMINDer* study was approved by the local Ethics Committee of the Technische Universität Dresden, Dresden, Germany (SR-EK-477112023, date of first approval: 2024-04-02). All procedures of the study will be carried out in compliance with the Declaration of Helsinki. Any substantial changes and additions to the study protocol will be submitted to the Ethics Committee of the Technische Universiät Dresden, Dresden as an amendment for review and approval. The study was registered on ClinicalTrials.gov (Identifier: NCT06530277, date of first submission: 2024-07-27) and adheres to Standard Protocol Items: Recommendations for Interventional Trials (SPIRIT) guidelines (99).

### 4.1 Informed Consent

Written informed consent for study procedures and data handling will be obtained from each participant. Prior to providing written consent, participants will be informed about the goals, content, and procedures of the *REMINDer* study, including the extent of measures that will be collected as well as the data management (see **section Study visits**). To ensure single blinding, participants will not be informed about the detailed primary and secondary outcomes. Participants will receive a compensation in return for their participation. In addition, participants receive a free intervention with the intervention costs being covered by the DZNE.

The DZNE is covered by a public liability insurance and has undertaken an accident insurance for the *REMINDer* study. Participants will be informed of this in the consent form prior to study participation. Participants will also be informed that they can withdraw at any time during the study, without any negative consequences. In case of withdrawal, participants can demand that their existing data will be completely deleted. A full disclosure of the study aims will be provided subsequent to the completion of the intervention study (after the t3 assessment) via written or oral debriefing.

### 4.2 Safety

The *REMINDer* study serves basic research purposes and will be based on voluntary participation. The study and implementation of the intervention are conducted according to the “do no harm” principle, meaning that the well-being and safety of participants are of highest priority. The sessions of the online intervention will be safe, as they are designed for the specific target group and will be applied by highly trained professionals. The facilitators/trainers will receive comprehensive training on the identification and mitigation of risks associated with the intervention and will collaborate with the study personnel to implement risk-mitigating procedures. During the online sessions, study personnel will be present to address individual needs of the participants, monitor the risk of injury, and provide technical support.

The *REMINDer* intervention is explicitly designed to promote positive effects on multiple health domains, including cognitive, psychological, physical, and social well-being in older adults. Eligibility criteria include potential physical constraints to ensure that safety is fulfilled throughout the study. In the case of concerns due to pre-existing subclinical health problems, participants will be required to provide a statement from their physician that they are able to participate in the study. Although we do everything in our power to minimize and avoid any potential harm, we cannot rule out the possibility of accidents or injuries occurring during the sessions. However, we believe that the risks of potential harm to participants are minimized.

### 4.3 Adverse event monitoring

The study follows the principles of good scientific / clinical practice and is considered to be safe for participants at any time point. Risks arising from participating in the study are considered to be low. Potential adverse events (AEs) and serious adverse events (SAEs) will be monitored throughout the *REMINDer* study. Participants will be informed to report AEs or SAEs in case of occurrence/notice. AEs and SAEs that occur during the intervention will be recorded by the study personnel in accordance with SOPs that have been established for this purpose. Participants will be given contact information in case of medical issues resulting from participation in the intervention. It is anticipated that no AEs/SAEs will result from participation in the *REMINDer* study. Written informed consent will be obtained from all participants to ensure that they are adequately informed about the safety standards in place.

### 4.4 Withdrawal or termination of study participation

Participants have the option to withdraw from the *REMINDer* study at any time point. The date of termination of study participation will be recorded in writing by the study personnel. In the event of early termination, participants will not be subject to any costs or disadvantages. There is no obligation for participants to provide a rationale for early termination.

## 5 Data collection, management, and dissemination

### 5.1 Data collection

During the *REMINDer* study, data pertaining to the examination of the participants, including personal information, will be collected and recorded at the DZNE Dresden. The data will be collected in either paper form or on electronic data carriers. The behavioural data will be collected via digital assessments (**see Figure 1**), of online questionnaires facilitated by the licensed LimeSurvey software. (https://www.limesurvey.org/). LimeSurvey is an established software utilized for the creation of online questionnaires and the collection of online data.

Participant data is collected in pseudonymized form (i.e., based on a non-personally identifiable number) on the LimeSurvey server and transferred to the secure DZNE server. The data will be managed by the study personnel via a registered LimeSurvey account. Access to this account will be restricted to study personnel. To ensure that questionnaires are correctly assigned to participants, each participant will be given a unique identification number, which will be numbered in ascending order for each participant. The identification number ensures the pseudonymization of the data.

### 5.2 Data management

The *REMINDer* study team will be responsible for data management under the guidance of the principal investigators (OK, MW). The data collected and stored in the study will be pseudonymized, that is, the inclusion of any identifying information such as names, initials, or dates of birth will be omitted. In addition, the data will be password protected within the participant database at DZNE Dresden; thus, the data will be secured against unauthorized access. An assignment of the pseudonymized data to a person is only possible via an identification list, which contains the participant’s pseudonymization (resp. identification) number and contact data (name, e-mail, telephone number). This identification list is stored separately from the study data at the DZNE Dresden and is only accessible to the study personnel. Ten years after the publication of the study, the identifying data will be deleted.

The data from the *REMINDer* study will be analysed in pseudonymized format by qualified scientific personnel according to the analysis strategy describe above (see **section Data analysis**). Publication of study results will not include any personal information and participants will not be identified. Given the sensitive and personal nature of the data, data from this study will not be publicly disclosed. Anonymized data will be made available to any qualified investigator upon request. The participants will be informed of their right to correct any data provided if necessary and to withdraw from the study at any time without consequence.

### 5.3 Dissemination of results

The results of the *REMINDer* study will be disseminated to the national and international scientific community, healthcare professionals, and other relevant stakeholders through publications in peer-reviewed journals and presentations at national and international scientific conferences. Publication of this study will be in accordance with the standards and recommendations of the international scientific community. The results will further be made available for both scientific and lay audiences on the ClinicalTrials.gov website. Additionally, the findings of this study will be disseminated to the general public and relevant stakeholders through media coverage including newspaper articles, social media platforms, and other dissemination channels.

## 6 Discussion

The *REMINDer* study constitutes an online randomized controlled pilot intervention study in older adults. The main objective of this study is to evaluate the feasibility and preliminary effectiveness of an online (home-based and live-streamed) multimodal mind-body group intervention to improve the mental and physical well-being of older participants compared to a passive control group (waitlist with delayed intervention). A sample of cognitively unimpaired older participants will be recruited from the general community in Germany and assessed before and after the intervention program using digital assessments with online questionnaires.

If successful, the present study may inform future trials and lifestyle-based strategies to impact mental health and well-being and other risk factors for dementia in older adults through online multimodal interventions. The online implementation and delivery of the *REMINDer* intervention is timely, cost-efficient, and provides a state-of-the-art mode of intervention, as shown by recent online randomized controlled studies (48, 49). The multimodal enrichment featured in our study integrates activities related to music, dance-based movement, and mindfulness in an interactive social environment that encourages social participation and integration. If shown to be feasible and effective, online multimodal mind-body interventions could serve as embodied prevention and intervention tools (13, 16) to help promote sustainable benefits for the health and well-being of older adults and prevent dementia, including AD, in the long term. In addition to the benefits for the individual, the intervention could also be important in reducing caregiver burden.

In general, environmental enrichment based on Arts for Health activities/interventions, such as dancing or playing music, are important for the integrated promotion of healthand well-being, as they can have positive impacts on multiple health domains, including mental, physical, and social health, and facilitate health-promoting behaviors (100). These intervention strategies have the potential to act as transdiagnostic tools, influencing health and well-being in heterogeneous older and younger populations. This may include healthy participants as well as patients with cognitive, mental, and physical conditions (22, 23, 101). Lifestyle-based intention strategies that enable early intervention and/or treatment of heterogeneous target groups could help to optimize available healthcare resources and address current gaps in prevention and care (102). The development and implementation of effective online multimodal mind-body interventions could represent be a significant step forward in reducing the economic, social, and emotional burden on individuals, caregivers, and healthcare providers.

## 7 Conclusions

To conclude, this randomized controlled study aims to provide evidence on the feasibility and impact of an online multimodal mind-body intervention delivered in an interactive group environment via videoconferencing. The active intervention will be compared to a passive control group (waitlist with delayed intervention). The *REMINDer* study may inform future trials and accessible lifestyle strategies to promote and maintain mental health benefits in older adults.

## Data Availability

Anonymized data will be made available to any qualified investigator upon request.

## 8 Acknowledgements

We would like to thank Clara Cornaro, Manuela Tobias, and Angela Nicotra, for their valuable expertise and their exceptional commitment and dedication to the training of the participants in the *REMINDer* study (C.C., M.T.) and the supervision of the facilitators (A.N.). We are very grateful to Karolin Schwellnus, Karla Paola Haug Reif, Adrianna Lipska-Dieck, Hanna Boscheck, Maxie Liebscher, Silke White and Amina Leib for their help and support with study preparation and/or participant recruitment and/or study conduct, including data collection, data management, and technical support. We would like to express our sincere gratitude to Dr. Theresa Köbe, Dr. Gerd Kempermann, and Dr. Peggy Looks for their intellectual input and/or expert scientific advice, and/or contribution to community outreach. Finally, we would like to thank all the participants in the *REMINDer* study.

This study received funding by the „*DZNE Stiftung – Forschung für ein Leben ohne Demenz, Parkinson und ALS“* (T 0531/36311/2020/kg and T 0531/41814/2002/kg). The funder was not involved in the study design, data acquisition, data analysis, data interpretation or manuscript writing.

## 9 Statements and declarations

Geben Sie hier eine Formel ein.

### 9.1 Authors contributions

M.W., S.S., O.P., A.M., S.K., O.K. Conceptualization, design, and methodology of the study; M.W., S.S., O.K. Implementation, administration, and conduct of the study including data collection; M.W., O.K. Supervision and funding acquisition; M.W., S.S., O.P., A.M., O.K. Writing-Original draft preparation, M.W., S.S., O.P., S.K., O.K. Writing-Reviewing and Editing

